# COVID-19: An Update on the Epidemiological, Genomic Origin, Phylogenetic study, India centric to Worldwide current status

**DOI:** 10.1101/2020.04.17.20070284

**Authors:** Murugan Nandagopal, R. Sagaya Jansi

**Author notes:** Corresponding Author: Dr Murugan. N M.Sc.,M.Phil., Ph.D., Scientist & Manager, Molecular Genetics, Lifecell International Pvt Ltd, chennai, India.

## Abstract

The pandemic spread of novel coronavirus, (SARS-CoV-2) causing CoronaVirus Infectious Diseases (COVID-19) emerged into a global threat for human life causing serious death rates and economic crunch all over the globe. As on April 17, 2020 at 2:00am CEST, there include a total of 2,034,802 confirmed cases for Corona and 1,35,163 deaths worldwide have been reported which includes 212 countries, areas or territories reported by World Health Organization (WHO), in which USA tops 6,32,781 confirmed cases (28,221 deaths) followed by Italy 1,65,155 (21,647 deaths), Spain 1,77,633 (18,579 deaths) and China 84,149 (4,642 deaths). This study aims to compare the genomic nature of SARS-CoV-2 genome reported from Wuhan, China with two Indian isolate genome reported by ICMR-NIV, India. Further Phylogenetic studies performed with coronavirus infecting non-human species like Bats, Duck, and sparrow were compared with Indian and other country whole genome sequences of SARS-CoV2 using MegaX and traced out the association between the human coronavirus with the other species viral genome. In addition, epidemiological reports on COVID-19 among Worldwide and India centric data were compared between April 7, 2020 to April 17, 2020 global data and the number of active cases were increased dramatically in this 10 days period studied, highlighted in the current study.

## Introduction

Totally 211 countries, territories or area have been affected by the novel pathogenic Human CoronaVirus (HCoV). The novel coronavirus was officially renamed as “SARS-CoV-2” from “2019-nCoV” by 11^th^ February 2020. The disease caused by SARS-CoV-2 was called “CoronaVirus Infectious Disease 2019” (COVID-19) by WHO. COVID-19 has been declared as a Public Health Emergency of International Concern by the WHO [1]. The virus belongs to Coronaviridae family which consists of single-stranded, non-segmented positive-sense RNA genome of size approximately 26-32 kb [2-3]. To date, six known HCoVs have been identified, namely HCoV-229E, HCoV-NL63, HCoV-OC43, HCoV-HKU1, Severe Acute Respiratory Syndrome Coronavirus (SARS-CoV) and Middle East respiratory syndrome coronavirus (MERS-CoV); except SARS and MERS, other four were universally dispersed in the human population and causes common cold infections among one-third of human populations [4-5].

Two serious coronavirus disease outbreaks occurred in the past two decades. First is SARS in 2003, originated from Southern China and spread to more than 30 countries and all major continents, resulted in more than 8000 human infections and 774 deaths [6-7]. The second include MERS in 2012 genetically different from SARS-CoV but infected 2,249 people in 27 countries with 35% case fatality [8-10].

The current COVID-19 entirely made the globe into lockdown status with the highest death rates and infected rates changing dynamically every day. The beginning of SARS-CoV-2 was linked to a cluster of patients with pneumonia of unknown aetiology connected to a local Huanan South China Seafood Market in Wuhan, Hubei Province, China in December 2019 [11]. It is now emerged as pandemic condition and World Health Organization (WHO) report as on April 17, 2020 at 2:00am CEST, there include a total of 2,034,802 confirmed cases for Corona and 1,35,163 deaths worldwide which includes 212 countries, areas or territories with cases [1] WHO region wise data in Fig.1A-B.

**Fig. 1A-B.**
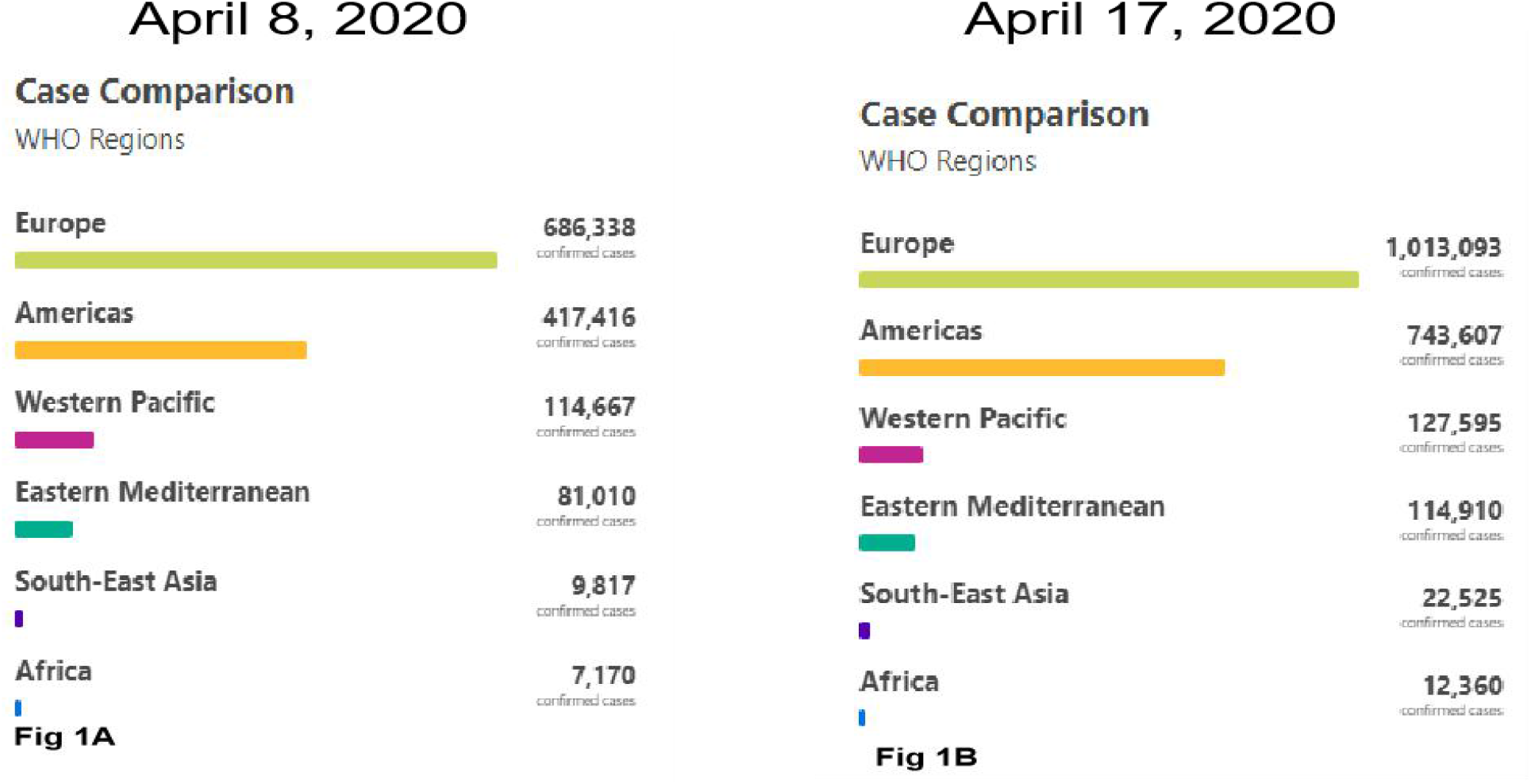
Region wise Confirmed cases as per WHO report as on (A) April 8, 2020 at 2:00am CET and (B) April 17, 2020 at 2:00 am CEST[1]

The fuss about the source of the virus and its intermediate host is still not yet confirmed. However, so far, there are still controversies about the source of the virus and its intermediate host [3]. The evolutionary analysis shows that the present strain of coronavirus is more similar to Bat coronavirus isolate RaTG13 (GenBank No.: MN996532.1), with 96.2% nucleotide homology in the whole genome [12-14]. Other groups suggest that pangolin, mink, snake, turtle may be potential intermediate hosts for the virus but not conclusively [3, 15-18].

In India, from January 25 to 31, 2020 three cases were shown to be positive for COVID-19 at Kerala state, all the three had travel history from Wuhan, China. Two patient’s samples were analysed by Next Generation sequencing and the data has been published in NCBI under MT012098.1 and MT050493.1 and GISAID database, accession numbers EPI ISL 413522 and EPI ISL 413523 and they reported briefly on various aspects [19]. We initiated our analysis, once the genome sequence was available at the NCBI database, however, Pragya D et al., reported by the end of March 2020. This study was aimed to understand the genomics and epidemiology of the Indian Human coronavirus with other major countries genome sequence, in addition the phylogeny relationship between COVID-19 from Indian patients genome with other Coronavirus genome which infects other species like Bats, Duck, Sparrow and etc.,

## Materials and Methods

### Sequence recovery and analysis

Genomic sequences hold innumerable information to foresee identification, location and pathogenicity of viral strains. As on April 07, 2020 there stay 547 nucleotide sequences and 5,364 protein sequences available in NCBI Virus database (https://www.nih.gov/coronavirus). Of which 469 sequences contains nearly about ∼28000-29000 base pairs (bps), sequences represents complete genome data and 78 denotes short specific protein encoding gene sequences.

The phylogeny was constructed to compare the evolutionary relationship of human and bat genome sequences. We used the first complete genome sequence submitted by China under the Genbank id: NC_045512.2 for Severe acute respiratory syndrome coronavirus 2 (SARS-CoV-2) [COVID-19] for the study and a BLAST similarity search was performed. The top 100 hits were screened and the aligned sequences were selected for further studies. Genome sequences of novel COVID-19 affecting Human (30) including 2 genome from India-Kerala state MT050493.1 & MT012098.1 and 2 closely related bat corona viral genomes MG772933.1 and MN996532.1 were investigated using MEGAX. The sequences were subjected to analysis by Maximum Likelihood method and Tamura-Nei model for the native and bootstrap tree (50 repeats) construction.

To compare the genomic evolutionary lineage from the other species, we compared the six (6) sequences of SARS-CoV-2 [Genbank id: NC_045512.2, MT126808.1, MT007544.1, MT049951.1, MT012098.1 (India) and MT050493.1 (India)] and 24 corona virus isolates from other species such as bat, sparrow, duck, pig, cow, etc., using MEGA X [20-22] (Table 1).

**Table 1.**
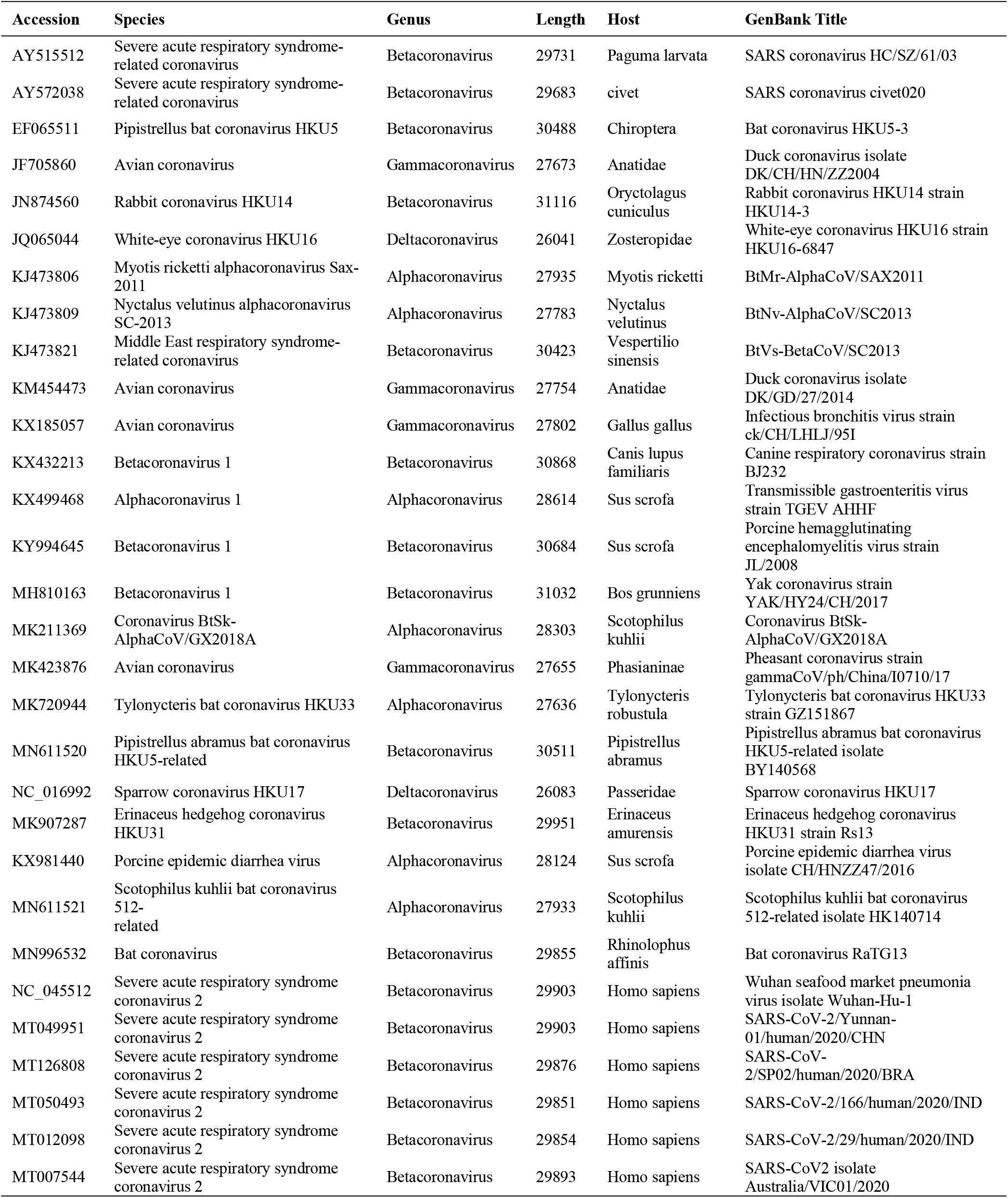
Genbank accession and details of Corona virus affecting other species genomes used in this study.

## Results

A phylogenetic tree was constructed with 30 genome sequences of Human coronavirus and 3 sequences of bat coronavirus using maximum likelihood method and Tamura-Nei model with MEGAX program. The algorithm produced a tree with highest log likelihood value (-632944.00) and also formed a bootstrap consensus tree inferred from 50 replicates. The percentage of trees in which the associated taxa clustered together was shown next to the branches. There were a total of 29903 positions in the final dataset. The lengths of the branches represent the amount of change that is estimated to have occurred between a pair of nodes. Alternative evolutionary trees are produced by bootstrapping analysis under maximum likelihood model. This is done through 50 iterations and when we recover the same node after 50 iterations then the node in the tree is well supported.

Upon boostrap, **Fig. 2**. data showed all the three Bat-SARS-like coronavirus in closed group branching to the China, USA and India strains. Though the first complete genome NC045512.2.1-29903 Wuhan-Hu-1 (blue triangle) was closely branched to MN996528.1 [Wuhan, Hubei, China], MT093571.1 [Swedan], MT039890.1 [Korean travelled from Wuhan, China], MT066156.1 [First case of Italy], MT007544.1 [Australia], the Bat SARS virus, precisely RaTG13 and SL-Covzc45 were completely close and neighborhood with the CoVid-19 isolated from Wuhan-genomes MT198652.2 (Spain), MT123292.2 (Guangzhou, China) MT012098.1 (Kerala, India) and LR757995.1 (Hong Kong, China).

**Fig. 2.**
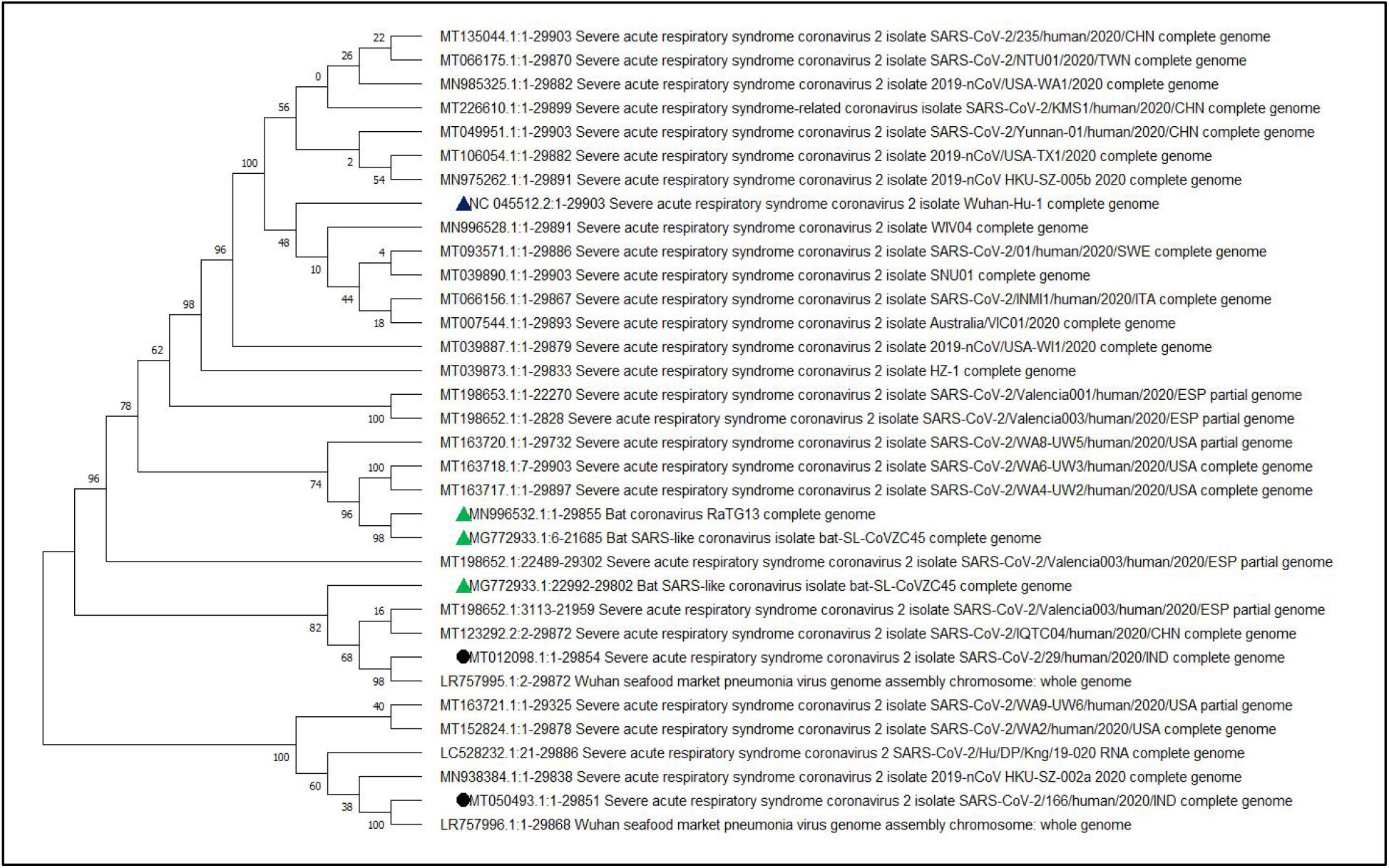
Phylogenetic tree (Bootstrap) of the complete genomes of severe acute respiratory syndrome coronavirus 2 viruses. Indian viruses are shown in black circle

The purpose of this study is to highlight the key role of Wuhan, China trace to Indian patients, Hence, the two India-Kerala state isolates were specifically traced by multiple alignments of the genome sequences using NCBI tool and Multalin, we found MT012098 consist of 29,854 bps and MT050493.1 consist of 29,851bps, three basepairs were missing in the initial sequence of the second isolate. Whereas totally, 9 point mutations were identified in this two genomes of which, 8 were C-T/T-C change and only one mutation at position 22,772 T/G change with 99% identity score, complete details recorded in the Table 2. The second strain from India MT050493.1 was in close association with LR757996.1 (Wuhan, China), MN938384.1 (Guangdong, China) and LC528232.1 (Isolated from a patient in cruise ship, Japan).

**Table 2:** Representation of the nucleotide position of both the Indian isolate genome and its nucleotide change

| Nucleotide Position | MT012098 | MT050493 |
| --- | --- | --- |
| 6488 | C | T |
| 6682 | T | C |
| 8769 | C | T |
| 14644 | T | C |
| 16864 | C | T |
| 17360 | T | C |
| 22772 | T | G |
| 24338 | C | T |
| 28131 | T | C |

Phylogenic study of coronavirus genome with diverse organisms showed, highest log likelihood value of -1064822.43 along with bootstrap consensus tree of 50 replicates. In this analysis a total of 31116 positions in the final dataset were used.

Though two Coronavirus from Indian patient had history from Wuhan travel and returned to India, Kerala state. In **Fig.3** human corona virus (MT012098.1) is closely related to White-eye coronavirus (JQ065044.1) and sparrow coronavirus (NC016992.1). The second Indian human coronavirus genome (MT050493.1) was closely related to Duck coronavirus (JF705860), Transmissible gastroenteritis (KX499468.1) and Canine respiratory coronavirus (KX432213.1). Whereas, all the other human affecting coronavirus genome from China, Wuhan, Brazile, Australia were closely related and they were adjacent to Bat coronavirus (MN996532.1, EF065511.1, MN611520.1), civet coronavirus AY572038.1 as a secondary neighborhood close to the human coronavirus next to bat virus. Though it is not conclusive, the coronavirus from sparrow and white-eye are more close to Indian isolates, which indicates they may be associated with the patients from other species of animal sources.

**Fig. 3.**
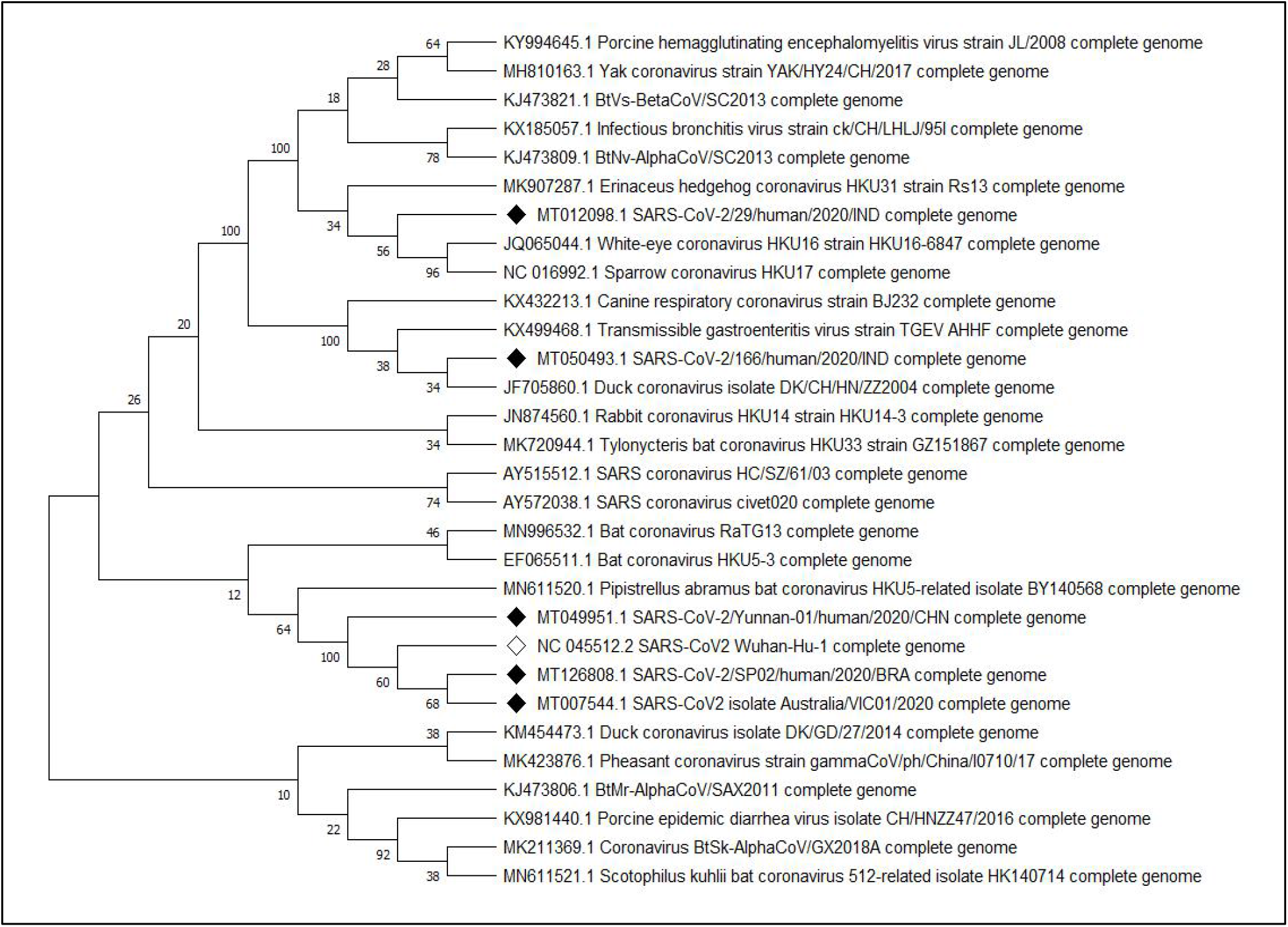
Phylogenetic tree (Bootstrap) of the complete genomes of Coronavirus from other species and human severe acute respiratory syndrome coronavirus 2 viruses. Indian viruses are shown in black diamond shape.

## Discussion

The emergence and rapid spread of COVID-19 signifies a perfect epidemiological storm. COVID-19 a member of the genus Betacoronavirus, subgenus - Sarbecovirus as like SARS-CoV, but MERS-CoV belongs to Merbecovirus (subgenus) [12, 23-24]. The RaTG13 is 96% identical to COVID-19 at the nucleotide level, but the receptor binding domain (RBD) is only 85% similar and shared only one of the six critical amino acid residues [25]. Still, the COVID-19 RBD for human ACE2 receptor found to be strong and functional [26] which armor the virus towards human respiratory system.

Research Group of South China Agricultural University analyzed more than 1,000 metagenomic samples, and they found 70% of pangolins were positive for the coronavirus. In addition, virus isolate from pangolin shared 99% sequence similarity with the current infected human strain SARS-CoV-2 [27] and they addressed pangolins may be one of the intermediate hosts for SARS-CoV-2 later they were embrassed to be a miscommunication of data reported [28] Infact, only 90% genome were homology to SARS-CoV-2 genome and Coronavirus from Pangolins. This incidence was a best example, how the scientific and public media were exaggerating the data without solid evident.

Inspite, the 96% identity of SARS-CoV-2 with the Bat coronavirus presumes, they are very closely related to SARSCoV-2, in virtual this likely represents more than 20 years of RNA sequence evolution, whcih may occure if the molecular clock emerges at an uncertain rate if there was strong adaptive evolution of the virus in humans[25]. Available bat coronavirus genome was studied from Yunnan province, over 1,500 km from Wuhan. There are few bat coronaviruses from Hubei province, and those that have been sequenced are relatively distant to SARS-CoV-2 in phylogenetic trees [29]. The available data on bat viruses is strongly biased toward some geographical locations, which needs further study [25].

The two Indian SARS-CoV-2 sequences were found to be non-identical (0.04% nt divergence), and the result of phylogenetic analysis indicated they are not in same clade, carries two different lineage which indicates they are non-identical source of infection [19]. Further multiple genome sequencing of Coronavirus from Indian patients will give us better understanding about the source of infection and mutation rate among the patient to patient and point of contact. The phylogenetic analysis of two Indian isolates elucidates strong associatedion with Bat virus genome, inspite the close association with duck and sparrow coronavirus were double checked with the genome blast given no statistical significance. Inspite, We estimate, if new coronavirus genome sequence from other animals like Bats, Pangolin, Civet or any other animal associated with Huanan South China Seafood Market in Wuhan, Hubei Province, China, may give us solid source of infection and the mode of transmission from animal to Human can be traced. Since, the 3-3.95 % genome difference in a RNA virus between SARS-CoV-2 and RaTG13 will not happen in a year and we are lacking the exact source or primary host of this novel coronavirus.

In India, as on April 17, 2020 18:00 IST, totally 13,815 confirmed cases, 1955 recovered cases, 11,403 active cases and 457 deaths. Currently, Maharastra, Delhi and Tamil Nadu were respectively top three affected States/Union Territory of India. The complete details were represented in the **Table 3**. Despite, India had cases from Wuhan to India-Kerala state by January 4^th^ week 2020, gradually; the cases were increased and identified from patients with travel history of various other countries and regions **Fig.4**.. The Government of India made complete Lockdown on March 24, 2020 midnight to April 15, 2020 early morning (21 days) as a first schedule. Second schedule announced by Honourable Prime Minister of India Mr. Narendra Modi on April 14^th^ 10 am, 2020 that the lockdown will be extended further to grab the spread of COVID-19 from local transmission (stage 2) to community transmission (stage 3) from April 15 early morning to May 3, 2020 early morning. This will further reduce the number of new infection rate, and the existing patients may be identified and treated according to the WHO guidelines. Inspite of all the effort taken, the rate of new cases has been steeply increased over the April 3^rd^ week and the list of confirmed, dead and recovered details shown in the Table.3.

**Fig. 4.**
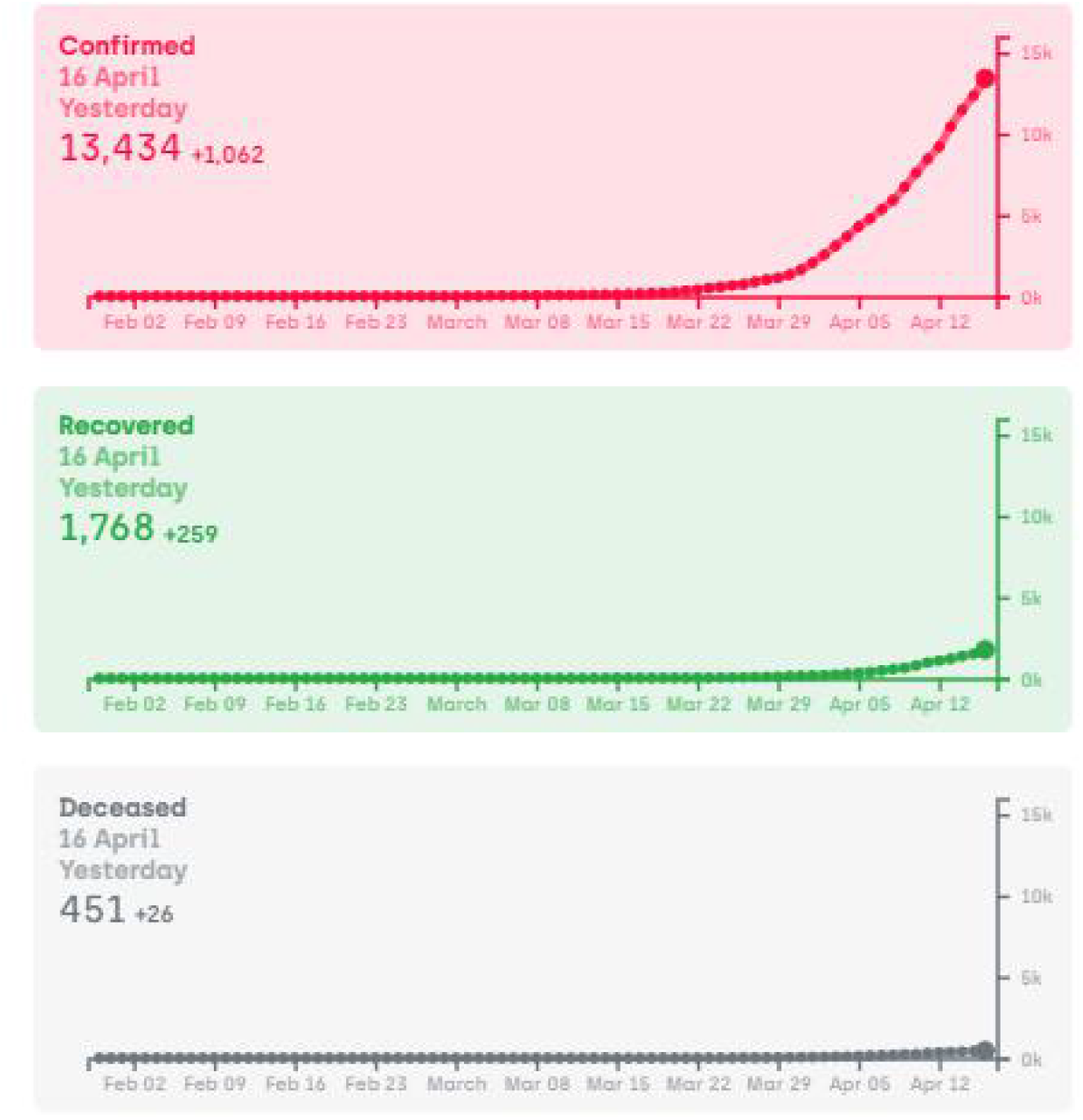
Graphical representation of confirmed, recovered, deceased cases from February 2020 to April 16, 2020 in India [Source :https://www.covid19india.org/]

**Table 3.** **Representation of State wise confirmed cases, Active cases, Recovered and Deceased cases as on April 17, 2020 18:55 IST** [Source :https://www.covid19india.org/]

| Position | STATE/Union Territory | CONFIRMED | ACTIVE | RECOVERED | DECEASED |
| --- | --- | --- | --- | --- | --- |
| 1 | Maharashtra | 3,202 | 2,708 | 300 | 194 |
| 2 | Delhi | 1,640 | 1,550 | 52 | 38 |
| 3 | Tamil Nadu | 1,323 | 1,025 | 283 | 15 |
| 4 | Rajasthan | 1,193 | 993 | 183 | 17 |
| 5 | Madhya Pradesh | 1,164 | 1,039 | 70 | 55 |
| 6 | Gujarat | 1,021 | 909 | 74 | 38 |
| 7 | Uttar Pradesh | 846 | 759 | 74 | 13 |
| 8 | Telangana | 700 | 495 | 187 | 18 |
| 9 | Andhra Pradesh | 572 | 523 | 35 | 14 |
| 10 | Kerala | 395 | 138 | 255 | 2 |
| 11 | Karnataka | 359 | 258 | 88 | 13 |
| 12 | Jammu and Kashmir | 314 | 271 | 38 | 5 |
| 13 | West Bengal | 255 | 194 | 51 | 10 |
| 14 | Haryana | 222 | 137 | 82 | 3 |
| 15 | Punjab | 211 | 167 | 30 | 14 |
| 16 | Bihar | 83 | 45 | 37 | 1 |
| 17 | Odisha | 60 | 40 | 19 | 1 |
| 18 | Uttarakhand | 40 | 31 | 9 | - |
| 19 | Chhattisgarh | 36 | 13 | 23 | - |
| 20 | Himachal Pradesh | 35 | 17 | 16 | 2 |
| 21 | Assam | 34 | 28 | 5 | 1 |
| 22 | Jharkhand | 29 | 27 | - | 2 |
| 23 | Chandigarh | 21 | 12 | 9 | - |
| 24 | Ladakh | 18 | 4 | 14 | - |
| 25 | Andaman and Nicobar Islands | 12 | 1 | 11 | - |
| 26 | Meghalaya | 9 | 8 | - | 1 |
| 27 | Goa | 7 | 1 | 6 | - |
| 28 | Puducherry | 7 | 6 | 1 | - |
| 29 | Manipur | 2 | 1 | 1 | - |
| 30 | Tripura | 2 | 1 | 1 | - |
| 31 | Arunachal Pradesh | 1 | - | 1 | - |
| 32 | Mizoram | 1 | 1 | - | - |
| 33 | Nagaland | 1 | 1 | - | - |
|  | Total | 13,815 | 11,403 | 1,955 | 457 |

Government of India and the health authorities of all the states had taken necessary action to curb spread and made containment plan and enhanced surveillances for the virus and its restricted movement. ICMR-NIV, Pune doing splendid work towards the identification of corona patients through Real-time quantitative PCR and rapid kit based methods. Currently, the diagnoses were carried out for the patients with clinical symptoms and or travel history or their contacts. In near future, the Government has planned to test everyone with symptoms irrespective of travel or contact history. This will curb the case of COVID-19 in India.

The world wide data alarms USA, Italy, Spain, UK, Iran, Netherlands and other countries with high rate of death cases, warrants early diagnosis and treatment, though there is no specific drug against COVID-19. The perfect lockdown, social distancing and safety measures plays vital role in this current scenario to reduce the number of new cases arises in the world Table 4 (7 parts). Fig 5A-C.

**Table 4:** Representative of complete country wise cumulative confirmed cases and cumulative deaths between April 7^th^ 2020 and April 17^th^ 2020 was calculated and the percentage wise confirmed cases and death rate in 10 days were shown in 7 parts (30 countries in each part), Totally 211 countries/areas or regions were given as per WHO report on April 17, 2020 **[1]**

| Position | Country Name | As on April 7, 2020 | As on April 17, 2020 | Confirmed cases in 10 days between 7.4.2020 to 17.4.2020 | Confirmed cases fold Increase in 10 days (Percentage) | As on April 7, 2020 | As on April 17, 2020 | Deaths in 10 days between 7.4.2020 to 17.4.2020 | Death fold Increase in 10 days (Percentage) |
| --- | --- | --- | --- | --- | --- | --- | --- | --- | --- |
|  |  | Cumulative Confirmed (numbers) | Cumulative Confirmed (numbers) |  |  | Cumulative Deaths (numbers) | Cumulative Deaths (numbers) |  |  |
| 1 | United States of America | 333811 | 632781 | 298970 | 47% | 9559 | 28221 | 18662 | 195% |
| 2 | Italy | 132547 | 165155 | 32608 | 20% | 16525 | 21647 | 5122 | 31% |
| 3 | Spain | 135032 | 177633 | 42601 | 24% | 13055 | 18579 | 5524 | 42% |
| 4 | France | 73488 | 105155 | 31667 | 30% | 8896 | 17146 | 8250 | 93% |
| 5 | The United Kingdom | 51612 | 98480 | 46868 | 48% | 5373 | 12868 | 7495 | 139% |
| 6 | Iran (Islamic Republic of) | 62589 | 77995 | 15406 | 20% | 3872 | 4869 | 997 | 26% |
| 7 | China | 83071 | 84149 | 1078 | 1% | 3340 | 4642 | 1302 | 39% |
| 8 | Belgium | 20814 | 33573 | 12759 | 38% | 1632 | 4440 | 2808 | 172% |
| 9 | Germany | 99225 | 130450 | 31225 | 24% | 1607 | 3569 | 1962 | 122% |
| 10 | Netherlands | 18803 | 28153 | 9350 | 33% | 1867 | 3134 | 1267 | 68% |
| 11 | Brazil | 11130 | 28320 | 17190 | 61% | 486 | 1736 | 1250 | 257% |
| 12 | Turkey | 30217 | 69392 | 39175 | 56% | 649 | 1518 | 869 | 134% |
| 13 | Sweden | 7206 | 11927 | 4721 | 40% | 477 | 1203 | 726 | 152% |
| 14 | Canada | 15806 | 28884 | 13078 | 45% | 293 | 1048 | 755 | 258% |
| 15 | Switzerland | 21574 | 26336 | 4762 | 18% | 641 | 973 | 332 | 52% |
| 16 | Portugal | 11730 | 18091 | 6361 | 35% | 311 | 599 | 288 | 93% |
| 17 | Indonesia | 2738 | 5516 | 2778 | 50% | 221 | 496 | 275 | 124% |
| 18 | Mexico | 2143 | 5847 | 3704 | 63% | 94 | 449 | 355 | 378% |
| 19 | Ireland | 5364 | 12547 | 7183 | 57% | 174 | 444 | 270 | 155% |
| 20 | India | 4421 | 12380 | 7959 | 64% | 114 | 414 | 300 | 263% |
| 21 | Ecuador | 3747 | 8225 | 4478 | 54% | 191 | 403 | 212 | 111% |
| 22 | Austria | 12297 | 14370 | 2073 | 14% | 220 | 393 | 173 | 79% |
| 23 | Romania | 4057 | 7216 | 3159 | 44% | 157 | 372 | 215 | 137% |
| 24 | Philippines | 3660 | 5660 | 2000 | 35% | 163 | 362 | 199 | 122% |
| 25 | Algeria | 1423 | 2268 | 845 | 37% | 173 | 348 | 175 | 101% |
| 26 | Denmark | 4681 | 6681 | 2000 | 30% | 187 | 309 | 122 | 65% |
| 27 | Poland | 4413 | 7582 | 3169 | 42% | 107 | 286 | 179 | 167% |
| 28 | Peru | 2281 | 11475 | 9194 | 80% | 83 | 254 | 171 | 206% |
| 29 | Russian Federation | 6343 | 27938 | 21595 | 77% | 47 | 232 | 185 | 394% |
| 30 | Republic of Korea | 10331 | 10635 | 304 | 3% | 192 | 230 | 38 | 20% |
|  |  | Cumulative Confirmed (numbers) | Cumulative Confirmed (numbers) |  |  | Cumulative Deaths (numbers) | Cumulative Deaths (numbers) |  |  |
| 31 | Dominican Republic | 1828 | 3755 | 1927 | 51% | 86 | 196 | 110 | 128% |
| 32 | Egypt | 1322 | 2505 | 1183 | 47% | 85 | 183 | 98 | 115% |
| 33 | Czechia | 4822 | 6303 | 1481 | 23% | 78 | 166 | 88 | 113% |
| 34 | Japan | 3906 | 9167 | 5261 | 57% | 80 | 148 | 68 | 85% |
| 35 | Hungary | 817 | 1652 | 835 | 51% | 47 | 142 | 95 | 202% |
| 36 | Colombia | 1485 | 3105 | 1620 | 52% | 35 | 131 | 96 | 274% |
| 37 | Norway | 5755 | 6677 | 922 | 14% | 59 | 130 | 71 | 120% |
| 38 | Morocco | 1141 | 2251 | 1110 | 49% | 83 | 128 | 45 | 54% |
| 39 | Pakistan | 4005 | 6919 | 2914 | 42% | 54 | 128 | 74 | 137% |
| 40 | Israel | 8611 | 12200 | 3589 | 29% | 52 | 126 | 74 | 142% |
| 41 | Argentina | 1554 | 2598 | 1044 | 40% | 46 | 115 | 69 | 150% |
| 42 | Ukraine | 1462 | 4162 | 2700 | 65% | 45 | 115 | 70 | 156% |
| 43 | Chile | 4815 | 8807 | 3992 | 45% | 37 | 105 | 68 | 184% |
| 44 | Panama | 1988 | 3751 | 1763 | 47% | 54 | 103 | 49 | 91% |
| 45 | Greece | 1755 | 2192 | 437 | 20% | 79 | 102 | 23 | 29% |
| 46 | Serbia | 2200 | 4873 | 2673 | 55% | 58 | 99 | 41 | 71% |
| 47 | Malaysia | 3793 | 5182 | 1389 | 27% | 62 | 84 | 22 | 35% |
| 48 | Saudi Arabia | 2795 | 6380 | 3585 | 56% | 41 | 83 | 42 | 102% |
| 49 | Iraq | 1031 | 1434 | 403 | 28% | 64 | 80 | 16 | 25% |
| 50 | Finland | 2176 | 3237 | 1061 | 33% | 28 | 72 | 44 | 157% |
| 51 | Luxembourg | 2843 | 3373 | 530 | 16% | 41 | 69 | 28 | 68% |
| 52 | Australia | 5844 | 6468 | 624 | 10% | 42 | 63 | 21 | 50% |
| 53 | Slovenia | 1021 | 1248 | 227 | 18% | 30 | 61 | 31 | 103% |
| 54 | Bangladesh | 164 | 1572 | 1408 | 90% | 17 | 60 | 43 | 253% |
| 55 | Puerto Rico | 513 | 1043 | 530 | 51% | 21 | 56 | 35 | 167% |
| 56 | South Africa | 1686 | 2605 | 919 | 35% | 12 | 48 | 36 | 300% |
| 57 | Republic of Moldova | 965 | 2049 | 1084 | 53% | 19 | 46 | 27 | 142% |
| 58 | Thailand | 2258 | 2672 | 414 | 15% | 27 | 46 | 19 | 70% |
| 59 | North Macedonia | 570 | 974 | 404 | 41% | 21 | 45 | 24 | 114% |
| 60 | Bosnia and Herzegovina | 695 | 1116 | 421 | 38% | 28 | 41 | 13 | 46% |
|  |  | Cumulative Confirmed (numbers) | Cumulative Confirmed (numbers) |  |  | Cumulative Deaths (numbers) | Cumulative Deaths (numbers) |  |  |
| 61 | Belarus | 700 | 3728 | 3028 | 81% | 13 | 36 | 23 | 177% |
| 62 | Bulgaria | 549 | 747 | 198 | 27% | 22 | 36 | 14 | 64% |
| 63 | San Marino | 277 | 393 | 116 | 30% | 32 | 36 | 4 | 13% |
| 64 | Estonia | 1108 | 1402 | 294 | 21% | 19 | 35 | 16 | 84% |
| 65 | Honduras | 298 | 426 | 128 | 30% | 22 | 35 | 13 | 59% |
| 66 | Tunisia | 596 | 780 | 184 | 24% | 22 | 35 | 13 | 59% |
| 67 | Croatia | 1222 | 1741 | 519 | 30% | 16 | 34 | 18 | 113% |
| 68 | Andorra | 540 | 673 | 133 | 20% | 21 | 33 | 12 | 57% |
| 69 | United Arab Emirates | 2076 | 5365 | 3289 | 61% | 11 | 33 | 22 | 200% |
| 70 | Burkina Faso | 345 | 543 | 198 | 36% | 17 | 32 | 15 | 88% |
| 71 | Afghanistan | 367 | 794 | 427 | 54% | 11 | 29 | 18 | 164% |
| 72 | Bolivia (Plurinational State of) | 183 | 441 | 258 | 59% | 11 | 29 | 18 | 164% |
| 73 | Lithuania | 843 | 1091 | 248 | 23% | 14 | 29 | 15 | 107% |
| 74 | Cuba | 350 | 862 | 512 | 59% | 9 | 27 | 18 | 200% |
| 75 | Albania | 377 | 494 | 117 | 24% | 22 | 25 | 3 | 14% |
| 76 | Democratic Republic of the Congo | 161 | 287 | 126 | 44% | 18 | 23 | 5 | 28% |
| 77 | Lebanon | 548 | 663 | 115 | 17% | 19 | 21 | 2 | 11% |
| 78 | Armenia | 833 | 1135 | 302 | 27% | 8 | 18 | 10 | 125% |
| 79 | Cameroon | 555 | 855 | 300 | 35% | 9 | 17 | 8 | 89% |
| 80 | Cyprus | 465 | 715 | 250 | 35% | 14 | 17 | 3 | 21% |
| 81 | Kazakhstan | 670 | 1295 | 625 | 48% | 6 | 16 | 10 | 167% |
| 82 | Niger | 253 | 609 | 356 | 58% | 10 | 15 | 5 | 50% |
| 83 | Azerbaijan | 641 | 1253 | 612 | 49% | 7 | 13 | 6 | 86% |
| 84 | undefined | 712 | 712 | 0 | 0% | 11 | 13 | 2 | 18% |
| 85 | Mali | 39 | 171 | 132 | 77% | 4 | 13 | 9 | 225% |
| 86 | Kenya | 142 | 234 | 92 | 39% | 4 | 11 | 7 | 175% |
| 87 | New Zealand | 943 | 1086 | 143 | 13% | 1 | 11 | 10 | 1000% |
| 88 | Nigeria | 232 | 373 | 141 | 38% | 5 | 11 | 6 | 120% |
| 89 | Singapore | 1375 | 4427 | 3052 | 69% | 6 | 10 | 4 | 67% |
| 90 | Kosovo[1] | 165 | 397 | 232 | 58% | 3 | 9 | 6 | 200% |

| Position | Country Name | As on April 7, 2020 | As on April 17, 2020 | Confirmed cases in 10 days between 7.4.2020 to | Cofirmed cases fold Increase in 10 days (Percentage) | As on April 7, 2020 | As on April 17, 2020 | Deaths in 10 days between 7.4.2020 to | Death fold Increase in 10 days (Percentage) |
| --- | --- | --- | --- | --- | --- | --- | --- | --- | --- |
|  |  | Cumulative Confirmed (numbers) | Cumulative Confirmed (numbers) |  |  | Cumulative Deaths (numbers) | Cumulative Deaths (numbers) |  |  |
| 91 | Mauritius | 244 | 324 | 80 | 25% | 7 | 9 | 2 | 29% |
| 92 | Sint Maarten | 37 | 57 | 20 | 35% | 6 | 9 | 3 | 50% |
| 93 | Uruguay | 406 | 493 | 87 | 18% | 6 | 9 | 3 | 50% |
| 94 | Venezuela (Bolivarian Republic of) | 159 | 197 | 38 | 19% | 3 | 9 | 6 | 200% |
| 95 | Bahamas | 29 | 53 | 24 | 45% | 5 | 8 | 3 | 60% |
| 96 | Ghana | 214 | 641 | 427 | 67% | 5 | 8 | 3 | 60% |
| 97 | Guadeloupe | 135 | 145 | 10 | 7% | 7 | 8 | 1 | 14% |
| 98 | Iceland | 1562 | 1727 | 165 | 10% | 6 | 8 | 2 | 33% |
| 99 | Martinique | 149 | 159 | 10 | 6% | 4 | 8 | 4 | 100% |
| 100 | Paraguay | 113 | 174 | 61 | 35% | 5 | 8 | 3 | 60% |
| 101 | Trinidad and Tobago | 105 | 114 | 9 | 8% | 8 | 8 | 0 | 0% |
| 102 | Bahrain | 811 | 1698 | 887 | 52% | 4 | 7 | 3 | 75% |
| 103 | Guernsey | 154 | 223 | 69 | 31% | 3 | 7 | 4 | 133% |
| 104 | Jordan | 349 | 401 | 52 | 13% | 6 | 7 | 1 | 17% |
| 105 | Qatar | 1832 | 4103 | 2271 | 55% | 4 | 7 | 3 | 75% |
| 106 | Sri Lanka | 180 | 238 | 58 | 24% | 6 | 7 | 1 | 17% |
| 107 | Côte d'Ivoire | 323 | 688 | 365 | 53% | 3 | 6 | 3 | 100% |
| 108 | El Salvador | 69 | 164 | 95 | 58% | 3 | 6 | 3 | 100% |
| 109 | Guyana | 29 | 55 | 26 | 47% | 4 | 6 | 2 | 50% |
| 110 | Jersey | 155 | 217 | 62 | 29% | 3 | 6 | 3 | 100% |
| 111 | Liberia | 14 | 73 | 59 | 81% | 3 | 6 | 3 | 100% |
| 112 | Slovakia | 534 | 863 | 329 | 38% | 2 | 6 | 4 | 200% |
| 113 | Barbados | 56 | 75 | 19 | 25% | 1 | 5 | 4 | 400% |
| 114 | Bermuda | 37 | 81 | 44 | 54% | 0 | 5 | 5 | 50% |
| 115 | Congo | 45 | 117 | 72 | 62% | 5 | 5 | 0 | 0% |
| 116 | Guam | 113 | 135 | 22 | 16% | 4 | 5 | 1 | 25% |
| 117 | Guatemala | 70 | 196 | 126 | 64% | 3 | 5 | 2 | 67% |
| 118 | Jamaica | 58 | 125 | 67 | 54% | 3 | 5 | 2 | 67% |
| 119 | Kyrgyzstan | 228 | 466 | 238 | 51% | 4 | 5 | 1 | 25% |
| 120 | Latvia | 542 | 666 | 124 | 19% | 1 | 5 | 4 | 400% |
|  |  | Cumulative Confirmed (numbers) | Cumulative Confirmed (numbers) |  |  | Cumulative Deaths (numbers) | Cumulative Deaths (numbers) |  |  |
| 121 | Somalia | 7 | 80 | 73 | 91% | 0 | 5 | 5 | 50% |
| 122 | Sudan | 14 | 32 | 18 | 56% | 2 | 5 | 3 | 150% |
| 123 | Togo | 44 | 81 | 37 | 46% | 3 | 5 | 2 | 67% |
| 124 | Costa Rica | 454 | 626 | 172 | 27% | 2 | 4 | 2 | 100% |
| 125 | Montenegro | 223 | 288 | 65 | 23% | 2 | 4 | 2 | 100% |
| 126 | Myanmar | 22 | 85 | 63 | 74% | 1 | 4 | 3 | 300% |
| 127 | Oman | 371 | 1019 | 648 | 64% | 2 | 4 | 2 | 100% |
| 128 | United Republic of Tanzania | 24 | 94 | 70 | 74% | 1 | 4 | 3 | 300% |
| 129 | Uzbekistan | 472 | 1349 | 877 | 65% | 2 | 4 | 2 | 100% |
| 130 | Ethiopia | 44 | 92 | 48 | 52% | 1 | 3 | 2 | 200% |
| 131 | Georgia | 195 | 336 | 141 | 42% | 2 | 3 | 1 | 50% |
| 132 | Haiti | 24 | 41 | 17 | 41% | 1 | 3 | 2 | 200% |
| 133 | Kuwait | 743 | 1524 | 781 | 51% | 1 | 3 | 2 | 200% |
| 134 | Malta | 241 | 399 | 158 | 40% | 0 | 3 | 3 | 30% |
| 135 | Mayotte | 164 | 233 | 69 | 30% | 2 | 3 | 1 | 50% |
| 136 | Zimbabwe | 9 | 23 | 14 | 61% | 1 | 3 | 2 | 200% |
| 137 | Angola | 16 | 19 | 3 | 16% | 2 | 2 | 0 | 0% |
| 138 | Antigua and Barbuda | 15 | 23 | 8 | 35% | 0 | 2 | 2 | 20% |
| 139 | Aruba | 64 | 95 | 31 | 33% | 0 | 2 | 2 | 20% |
| 140 | Belize | 7 | 18 | 11 | 61% | 1 | 2 | 1 | 100% |
| 141 | Djibouti | 121 | 591 | 470 | 80% | 0 | 2 | 2 | 20% |
| 142 | Isle of Man | 127 | 254 | 127 | 50% | 1 | 2 | 1 | 100% |
| 143 | Malawi | 4 | 16 | 12 | 75% | 0 | 2 | 2 | 20% |
| 144 | Northern Mariana Islands (Commonwealth of the) | 8 | 13 | 5 | 38% | 1 | 2 | 1 | 100% |
| 145 | occupied Palestinian territory, including east Jerusalem | 254 | 294 | 40 | 14% | 1 | 2 | 1 | 100% |
| 146 | Saint Martin | 31 | 35 | 4 | 11% | 2 | 2 | 0 | 0% |
| 147 | Senegal | 226 | 335 | 109 | 33% | 2 | 2 | 0 | 0% |
| 148 | Syrian Arab Republic | 19 | 33 | 14 | 42% | 2 | 2 | 0 | 0% |
| 149 | Zambia | 39 | 48 | 9 | 19% | 1 | 2 | 1 | 0% |
| 150 | Benin | 23 | 37 | 14 | 38% | 1 | 1 | 0 | 0% |

| Position | Country Name | As on April 7, 2020 | As on April 17, 2020 | Confirmed cases in 10 days between 7.4.2020 to 17.4.2020 | Cofirmed cases fold Increase in 10 days (Percentage) | As on April 7, 2020 | As on April 17, 2020 | Deaths in 10 days between 7.4.2020 to 17.4.2020 | Death fold Increase in 10 days (Percentage) |
| --- | --- | --- | --- | --- | --- | --- | --- | --- | --- |
|  |  | Cumulative Confirmed (numbers) | Cumulative Confirmed (numbers) |  |  | Cumulative Deaths (numbers) | Cumulative Deaths (numbers) |  |  |
| 151 | Botswana | 6 | 15 | 9 | 60% | 1 | 1 | 0 | 0% |
| 152 | Brunei Darussalam | 135 | 136 | 1 | 1% | 1 | 1 | 0 | 0% |
| 153 | Cabo Verde | 7 | 55 | 48 | 87% | 1 | 1 | 0 | 0% |
| 154 | Cayman Islands | 39 | 60 | 21 | 35% | 1 | 1 | 0 | 0% |
| 155 | Curacao | 13 | 14 | 1 | 7% | 1 | 1 | 0 | 0% |
| 156 | Eswatini | 9 | 16 | 7 | 44% | 0 | 1 | 1 | 0% |
| 157 | Gabon | 21 | 95 | 74 | 78% | 1 | 1 | 0 | 0% |
| 158 | Gambia | 4 | 9 | 5 | 56% | 1 | 1 | 0 | 0% |
| 159 | Guinea | 111 | 438 | 327 | 75% | 0 | 1 | 1 | 0% |
| 160 | Libya | 19 | 48 | 29 | 60% | 1 | 1 | 0 | 0% |
| 161 | Liechtenstein | 78 | 81 | 3 | 4% | 1 | 1 | 0 | 0% |
| 162 | Mauritania | 6 | 7 | 1 | 14% | 1 | 1 | 0 | 0% |
| 163 | Nicaragua | 6 | 9 | 3 | 33% | 1 | 1 | 0 | 0% |
| 164 | Suriname | 10 | 10 | 0 | 0% | 1 | 1 | 0 | 0% |
| 165 | Turks and Caicos Islands | 5 | 11 | 6 | 55% | 1 | 1 | 0 | 0% |
| 166 | United States Virgin Islands | 43 | 51 | 8 | 16% | 1 | 1 | 0 | 0% |
| 167 | Anguilla | 3 | 3 | 0 | 0% | 0 | 0 | 0 | 0% |
| 168 | Bhutan | 5 | 5 | 0 | 0% | 0 | 0 | 0 | 0% |
| 169 | Bonaire, Sint Eustatius and Saba | 2 | 4 | 2 | 50% | 0 | 0 | 0 | 0% |
| 170 | British Virgin Islands | 3 | 3 | 0 | 0% | 0 | 0 | 0 | 0% |
| 171 | Burundi | 3 | 5 | 2 | 40% | 0 | 0 | 0 | 0% |
| 172 | Cambodia | 115 | 122 | 7 | 6% | 0 | 0 | 0 | 0% |
| 173 | Central African Republic | 9 | 12 | 3 | 25% | 0 | 0 | 0 | 0% |
| 174 | Chad | 9 | 27 | 18 | 67% | 0 | 0 | 0 | 0% |
| 175 | Dominica | 14 | 16 | 2 | 13% | 0 | 0 | 0 | 0% |
| 176 | Equatorial Guinea | 16 | 51 | 35 | 69% | 0 | 0 | 0 | 0% |
| 177 | Eritrea | 29 | 35 | 6 | 17% | 0 | 0 | 0 | 0% |
| 178 | Falkland Islands (Malvinas) | 2 | 11 | 9 | 82% | 0 | 0 | 0 | 0% |
| 179 | Faroe Islands | 181 | 184 | 3 | 2% | 0 | 0 | 0 | 0% |
| 180 | Fiji | 15 | 17 | 2 | 12% | 0 | 0 | 0 | 0% |
|  |  | Cumulative Confirmed (numbers) | Cumulative Confirmed (numbers) |  |  | Cumulative Deaths (numbers) | Cumulative Deaths (numbers) |  |  |
| 181 | French Guiana | 68 | 96 | 28 | 29% | 0 | 0 | 0 | 0% |
| 182 | French Polynesia | 42 | 55 | 13 | 24% | 0 | 0 | 0 | 0% |
| 183 | Gibraltar | 103 | 129 | 26 | 20% | 0 | 0 | 0 | 0% |
| 184 | Greenland | 11 | 11 | 0 | 0% | 0 | 0 | 0 | 0% |
| 185 | Grenada | 12 | 14 | 2 | 14% | 0 | 0 | 0 | 0% |
| 186 | Guinea-Bissau | 33 | 46 | 13 | 28% | 0 | 0 | 0 | 0% |
| 187 | Holy See | 7 | 8 | 1 | 13% | 0 | 0 | 0 | 0% |
| 188 | Lao People's Democratic Republic | 12 | 19 | 7 | 37% | 0 | 0 | 0 | 0% |
| 189 | Madagascar | 85 | 117 | 32 | 27% | 0 | 0 | 0 | 0% |
| 190 | Maldives | 19 | 23 | 4 | 17% | 0 | 0 | 0 | 0% |
| 191 | Monaco | 40 | 93 | 53 | 57% | 0 | 0 | 0 | 0% |
| 192 | Mongolia | 15 | 31 | 16 | 52% | 0 | 0 | 0 | 0% |
| 193 | Montserrat | 6 | 11 | 5 | 45% | 0 | 0 | 0 | 0% |
| 194 | Mozambique | 10 | 29 | 19 | 66% | 0 | 0 | 0 | 0% |
| 195 | Namibia | 16 | 16 | 0 | 0% | 0 | 0 | 0 | 0% |
| 196 | Nepal | 9 | 16 | 7 | 44% | 0 | 0 | 0 | 0% |
| 197 | New Caledonia | 18 | 18 | 0 | 0% | 0 | 0 | 0 | 0% |
| 198 | Papua New Guinea | 1 | 7 | 6 | 86% | 0 | 0 | 0 | 0% |
| 199 | Réunion | 349 | 394 | 45 | 11% | 0 | 0 | 0 | 0% |
| 200 | Rwanda | 104 | 138 | 34 | 25% | 0 | 0 | 0 | 0% |
| 201 | Saint Barthélemy | 6 | 6 | 0 | 0% | 0 | 0 | 0 | 0% |
| 202 | Saint Kitts and Nevis | 10 | 14 | 4 | 29% | 0 | 0 | 0 | 0% |
| 203 | Saint Lucia | 14 | 15 | 1 | 7% | 0 | 0 | 0 | 0% |
| 204 | Saint Vincent and the Grenadines | 7 | 12 | 5 | 42% | 0 | 0 | 0 | 0% |
| 205 | Sao Tome and Principe | 4 | 4 | 0 | 0% | 0 | 0 | 0 | 0% |
| 206 | Seychelles | 11 | 11 | 0 | 0% | 0 | 0 | 0 | 0% |
| 207 | Sierra Leone | 6 | 15 | 9 | 60% | 0 | 0 | 0 | 0% |
| 208 | South Sudan | 1 | 4 | 3 | 75% | 0 | 0 | 0 | 0% |
| 209 | Timor-Leste | 1 | 18 | 17 | 94% | 0 | 0 | 0 | 0% |
| 210 | Uganda | 52 | 55 | 3 | 5% | 0 | 0 | 0 | 0% |
| 211 | Viet Nam | 245 | 268 | 23 | 9% | 0 | 0 | 0 | 0% |

**Fig. 5A-C.**
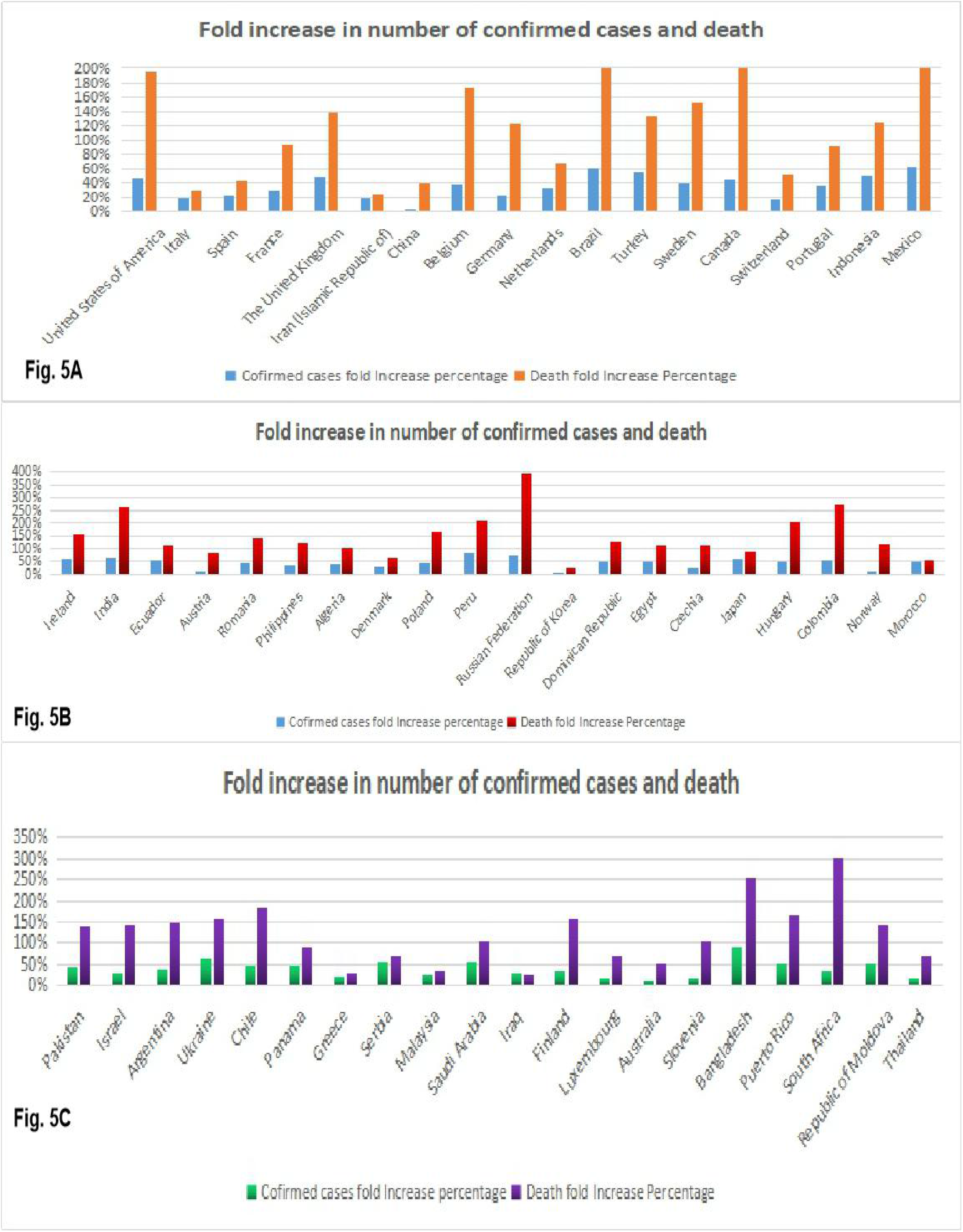
Representation of major country wise confirmed case fold change and Death fold change between April 7, 2020 to April 17, 2020 (10 days) as per WHO report [1].

Inspite of all the effort taken all over the world, USA has been affected to the highest level in the world, where 6,32,781 confirmed cases and 28,221deaths were reported as on April 17, 2020. Next to USA, the top 9 countries were Italy, Spain, France, The United Kingdom, Iran (Islamic Republic of), China, Belgium, Germany, Netherlands have confirmed cases of 1,65,155, 1,77,633, 1,05,155, 98,480, 77,995, 84,149, 33,573, 1,30,450 and 2,81,53 respectively and the death toll reported to be Italy 21,647, Spain 18,579, France 17,146, The United Kingdom 12,868, Iran (Islamic Republic of) 4,869, China 4,642, Belgium 4,440, Germany 3,569, Netherlands 3,134 as per WHO report (April 17,2020 2:00am CEST). Although Germany position to be 4^th^ as per the confirmed cases, but the number of death reported till date was lower compared to other countries, which made it as 9^th^ position. The complete country wise cumulative confirmed cases and cumulative deaths between April 7^th^ 2020 and April 17^th^ 2020 was calculated and the percentage wise confirmed cases and death rate in 10 days was calculated and shown in the Table. 4.

The percentage of death rate increased in just 10 days alarms the world and it should be recorded in the literature for future study. Hence, we included the whole data in the table, and the top 20 countries with death fold increase in 10 days between April 7, 2020 to April 17, 2020 as follows United States of America 195%, Italy 31%, Spain 42%, France 93%, The United Kingdom 139%, Iran (Islamic Republic of) 26%, China 39%, Belgium 172%, Germany 122%, Netherlands 68%, Brazil 257%, Turkey 134%, Sweden 152%, Canada 258%, Switzerland 52%, Portugal 93%, Indonesia 124%, Mexico 378%, Ireland 155%, India 263%.

Continued testing of people, irrespective of symptoms and travel history is the need of time to scrutinize the asymptomatic carrier people among the group of population, will cut down the number of death rate. This April 2020 plays major role in the steady death toll increase among various countries mentioned above. Early diagnosis, appropriate treatment, containment zone formation, Quarantine the people with symptoms may help us to flat the rate of infection and death rate.

### Possible ending of COVID-19

The pandemic COVID-19 outbreak may have handful different endings. If nature or God give us the luck, the most favourable scenario would be COVID-19 unconsciously petering out as was the case with SARS in 2003. The second chance would be like MERS which, continue sporadically pop up over the years. The third one in worst scenario, it may create more deaths over the year as the sinister path like 1918 Spanish influenza over a decade [30]. Let the nature and future determine it.

## Conclusion

The dynamic increase of number of confirmed cases and genome sequences from all over the world made us to register the various conditions as on April 1^st^ week, 2020. Every one hour, the world wide data has been updated in various resources like WHO, John Hopkins website, Worldometer. In this context, our study from 2 Indian isolate provided us the trace for infection source and the spread rate increases the point mutation, which may either reduce the virulent or increase the pathogenicity of novel coronavirus, SARS-CoV-2 mediated COVID-19 which needs further study.

## Data Availability

Data available from public source- NCBI

https://www.ncbi.nlm.nih.gov/labs/virus/vssi/#/virus?SeqType_s=Nucleotide&VirusLineage_ss=Wuhan%20seafood%20market%20pneumonia%20virus,%20taxid:2697049&utm_campaign=wuhan_nCoV&utm_source=insights&utm_medium=referral

## Acknowledgement

I thank **Dr Giridharan Appaswamy**, Chief Scientific Officer, LifeCell International Private Limited, Chennai, for his continuous support and encouragement to do this research work. I thank Management, Lifecell International Pvt Ltd and all the lab members from Lifecell, R& D division for their support during this work.

